# Visual evoked potential in myelin oligodendrocyte glycoprotein antibody-associated disease

**DOI:** 10.1101/2024.07.22.24310743

**Authors:** Yano Chikashi, Matsuura Eiji, Nakamura Tomonori, Sonoda Ayako, Shigehisa Ayano, Ando Masahiro, Nozuma Satoshi, Higuchi Yujiro, Sakiyama Yusuke, Hashiguchi Akihiro, Michizono Kumiko, Takashima Hiroshi

**Affiliations:** Department of Neurology and Geriatrics, Kagoshima University Graduate School of Medical and Dental Sciences, 8-35-1 Sakuragaoka, Kagoshima 890-8520, Japan

**Keywords:** Optic Neuritis, Myelin-Oligodendrocyte Glycoprotein associated disease, Visual Evoked Potentials, Neuromyelitis Optica, Multiple Sclerosis, Diagnostic Techniques

## Abstract

The visual evoked potential (VEP) patterns of optic neuritis are known to often differ between multiple sclerosis (MS) and neuromyelitis optica spectrum disorder (NMOSD) but have been less reported in myelin oligodendrocyte glycoprotein antibody-associated disease (MOGAD). This study aimed to characterize the VEP pattern in MOGAD and evaluate its utility in distinguishing MOGAD from MS and NMOSD. We retrospectively reviewed the clinical manifestations and VEP findings in patients with MS (n = 29), NMOSD (n = 14), and MOGAD (n = 10). In eyes with acute visual impairment, VEP responses were detectable in all eyes with MOGAD but were undetectable in a significant percentage of eyes with MS (27.3%) and NMOSD (42.9%). In addition, VEP abnormalities in eyes without acute visual impairment were rare in MOGAD (23.1%) compared to MS (55.3%) and NMOSD (42.9%). Our results indicated that subclinical VEP abnormalities or undetectable VEP responses were less common in patients with MOGAD compared to patients with MS and NMOSD. VEP testing demonstrates potential diagnostic utility in distinguishing among these conditions.

## 1. Introduction

Myelin oligodendrocyte glycoprotein antibody-associated disease (MOGAD) is an inflammatory demyelinating disorder of the central nervous system and targets the myelin oligodendrocyte glycoprotein (MOG), which is an essential constituent in myelin sheath formation (Johns and Bernard, 1999). MOGAD exhibits a diverse clinical spectrum, including optic neuritis (ON), myelitis, cortical encephalitis, acute disseminated encephalomyelitis, and brainstem encephalitis. MOG antibodies are notably prevalent in pediatric cases of acute disseminated encephalomyelitis and are present in pediatric and adult ON (Banwell et al., 2023). Subsequently, case reports of MOG antibody-positive cases in individuals previously diagnosed with aquaporin-4 (AQP-4) antibody-negative neuromyelitis optica spectrum disorder (NMOSD) have emerged (Kitley et al., 2012). Remarkably, ON is often the initial presenting symptom of adult-onset MOGAD (Banwell et al., 2023). The optic nerve lesions often extend bilaterally and longitudinally and are localized anterior to the optic nerve and optic disc. In multiple sclerosis (MS), optic nerve lesions tend to be unilateral and are localized anterior to the optic nerve (Carnero Contentti et al., 2023). NMOSD is an antibody-mediated inflammatory disorder that targets AQP-4, which is a protein expressed in astrocytes. Notably, NMOSD-associated ON frequently extends bilaterally across the optic chiasm and often results in a severe disability that affects both eyes (Carnero Contentti et al., 2023; Merle et al., 2007). These three diseases present with different symptoms and respond differently to treatment. Nevertheless, no method other than antibody testing and MRI has been reported to effectively distinguish between these diseases. Thus, in cases where antibody testing yields negative results, differentiating these three diseases becomes challenging.

Visual evoked potential (VEP) is commonly used to assess ON by capturing the electrical signals that are generated in the visual cortex; originate from the retina; and traverse the optic nerve, optic chiasm, optic tract, and lateral geniculate body, then ultimately reaching the occipital lobe. Recording electrical potentials on the scalp over the occipital lobe enables noninvasive and individualized evaluation of the functionality of each optic nerve (Odom et al., 2016). The distinctive VEP patterns in MS and NMOSD and their utility in distinguishing between the two conditions are well documented (Neto et al., 2013). In MS, P100 latency is frequently prolonged, even in clinically unaffected eyes or during periods without visual abnormalities (Grecescu, 2014; Sisto et al., 2005). NMOSD often presents with reduced P100 amplitude or undetectable VEP responses. These features indicate that pathological abnormalities can affect VEP outcomes (Shen et al., 2019). Conversely, there had been limited available literature on the VEP patterns in MOGAD, and comparative studies with MS and NMOSD remain scarce.

The objectives of this study were to review the clinical and electrophysiological features and distinctive VEP patterns of ON in MOGAD and to evaluate the utility of VEP in distinguishing MOGAD from MS and NMOSD.

## 2. Materials and Methods

### 2.1. Study population

We retrospectively collected the data of patients with MS, NMOSD, and MOGAD who underwent VEP testing at the Neurology Department of Kagoshima University Hospital, Japan between January 2015 and May 2024. The diagnoses of MS, NMOSD, and MOGAD were based on the respective diagnostic criteria in 2017, 2015, and 2023 (Banwell et al., 2023; Thompson et al., 2018; Wingerchuk et al., 2015). We excluded patients in whom the specific antibody markers for NMOSD and MOGAD were absent in order to minimize selection bias. MOG-antibody was evaluated with the live cell-based assay using cells expressing full-length MOG. The single-facility setting ensured uniform data collection and reduced protocol-related bias. The study protocol was approved by the institutional review board of Kagoshima University (approval number #230141).

### 2.2. Data extraction

The collected data included the age at the time of VEP testing; age at disease onset; sex; presence or absence of self-reported visual impairment at the time of VEP testing; prior history of visual impairment; VEP parameters; Expanded Disability Status Scale (EDSS); visual acuity; and the presence of lesions in the brain, brainstem, and spinal cord. Considering that the average hospitalization period for patients with MS, NMOSD, and MOGAD was approximately one month at our institution, eyes with acute phase ON were defined as those with VEP testing performed within 40 days of self-reported visual impairment onset, and eyes with prior ON were defined as those with VEP testing performed more than 40 days after onset. If multiple examinations were conducted during this period, the data from the initial VEP testing after the onset of visual impairment were included. For patients who had no visual impairment, the initial VEP testing results were included. In cases of undetectable VEP responses, the amplitude was treated as 0, and latency was not evaluated.

### 2.3. Pattern-reversal visual evoked potential

Neuropack X1 (Nihon Kohden, Tokyo, Japan) was used for recording VEP. For full-field pattern-reversal VEP, we used a 17-inch liquid crystal display monitor, which was positioned at the eye level and maintained at a constant distance of 121 cm from the patient. The stimulus reversal frequency was set to 1 Hz, and the check size was fixed at 32, with a visual field angle of 0.5°. The filter frequency range was 1–200 Hz.

Electrode placement followed the international 10/20 system. The active electrode was positioned at Oz, the reference electrode was placed at Fz, and the ground electrode was located on the wrist. The recording montage featured Oz-Fz configurations. Data recording involved the acquisition of 200 consecutive averages that were triggered by each pattern-reversal event (Odom et al., 2016). Full field monocular stimulation was administered separately to the left and right eyes. The gaze of the participants remained steadily fixed on the center of the monitor, with vigilance and sustained attention ensured throughout the recording process.

Full-field stimulation was individually performed for each eye, and the P100 peak latency at Oz-Fz was included in this study. At our institution, prolonged P100 latency was defined as exceeding the upper limit of the reference value for monocular P100 peak latency, which was set at 126 ms (mean + 2 × standard deviation). P100 amplitude was measured between the N75 and P100 peaks. However, specific reference values for P100 amplitude have not been established. Reference values were calculated based on VEP test results from 10 healthy individuals, comprised of 5 women and 5 men, with a median age of 30 years (interquartile range [IQR], 28.3–33 years) (control group).

### 2.4. Statistical analysis

Results were presented as median with IQR. Wilcoxon rank-sum test was employed to compare the differences in P100 latency and P100 amplitude between eyes affected by ON and normal eyes in MS, NMOSD, and MOGAD groups. Statistical significance was designated as *p <* 0.05. Statistical analysis was conducted using RStudio (version 4.2.1).

## 3. Results

### 3.1. Multiple sclerosis group

The MS group comprised a total of 29 patients, who had a median age at disease onset of 32 years (IQR, 26–40 years). The median age at the time of VEP testing was 36 years (IQR, 28–45 years), and the disease duration ranged from 7 days to 16 years, with a median of 3 months (IQR, 1–76 months). At the time of VEP testing, the patients’ EDSS median score was 3 (IQR, 2-5). Visual acuity assessments were available for 17 of 29 patients, with a median of 1.0.

Of the 29 patients, 27 had cerebral lesions, 13 had brainstem lesions, and 20 had spinal cord lesions (Table S1). VEP testing was performed in 9 patients (4 women and 5 men) with acute phase ON and in 20 patients (12 women and 8 men) without acute phase ON, including those with a prior history of ON or without a history of ON (Table 1).

**Table 1.** Clinical features in MS, NMOSD, and MOGAD.

|  | MS |  | NMOSD |  | MOGAD |  |
| --- | --- | --- | --- | --- | --- | --- |
| Optic neuritis | ON (+) | ON (-) | ON (+) | ON (-) | ON (+) | ON (-) |
| Number of patients | 9 | 20 | 7 | 7 | 5 | 5 |
| Female/male | 4/5 | 12/8 | 6/1 | 4/3 | 3/2 | 3/2 |
| Age at VEP study, year, median (IQR) | 35 (25–45) | 33 (27.5–45) | 49 (40–56.5) | 59 (50.5–66) | 31 (29–45) | 37 (32–44) |
| EDSS, median (IQR) | 3.0 (2-5) | 3.25 (2-4.625) | 5 (3.25-5) | 4.0 (2.75-5.75) | 2.0 (1.5-3.0) | 4.0 (4-4.5) |

MS: multiple sclerosis, NMOSD: neuromyelitis optica spectrum disorder, MOGAD: myelin oligodendrocyte glycoprotein antibody-associated disease, ON: acute phase optic neuritis, VEP: visual evoked potential, IQR: interquartile range. EDSS: extended disability status scale.

Among the MS cases, acute phase ON was seen in 11 eyes (bilateral in 2 cases and unilateral in 7 cases). Of the 11 eyes with acute phase ON, 5 had prolonged P100 latency, and 3 had undetectable VEP responses. Of the remaining 47 eyes without acute phase ON, including 7 eyes with a prior history of ON and 40 eyes without a history of ON, 21 had prolonged P100 latency, and 5 had undetectable VEP responses (Table 2, 3). There were no significant differences in P100 latency and amplitude between the eyes with acute phase ON and the other eyes (*p* = 0.37 and *p* = 0.70, respectively) (Figure 1).

**Table 2.**
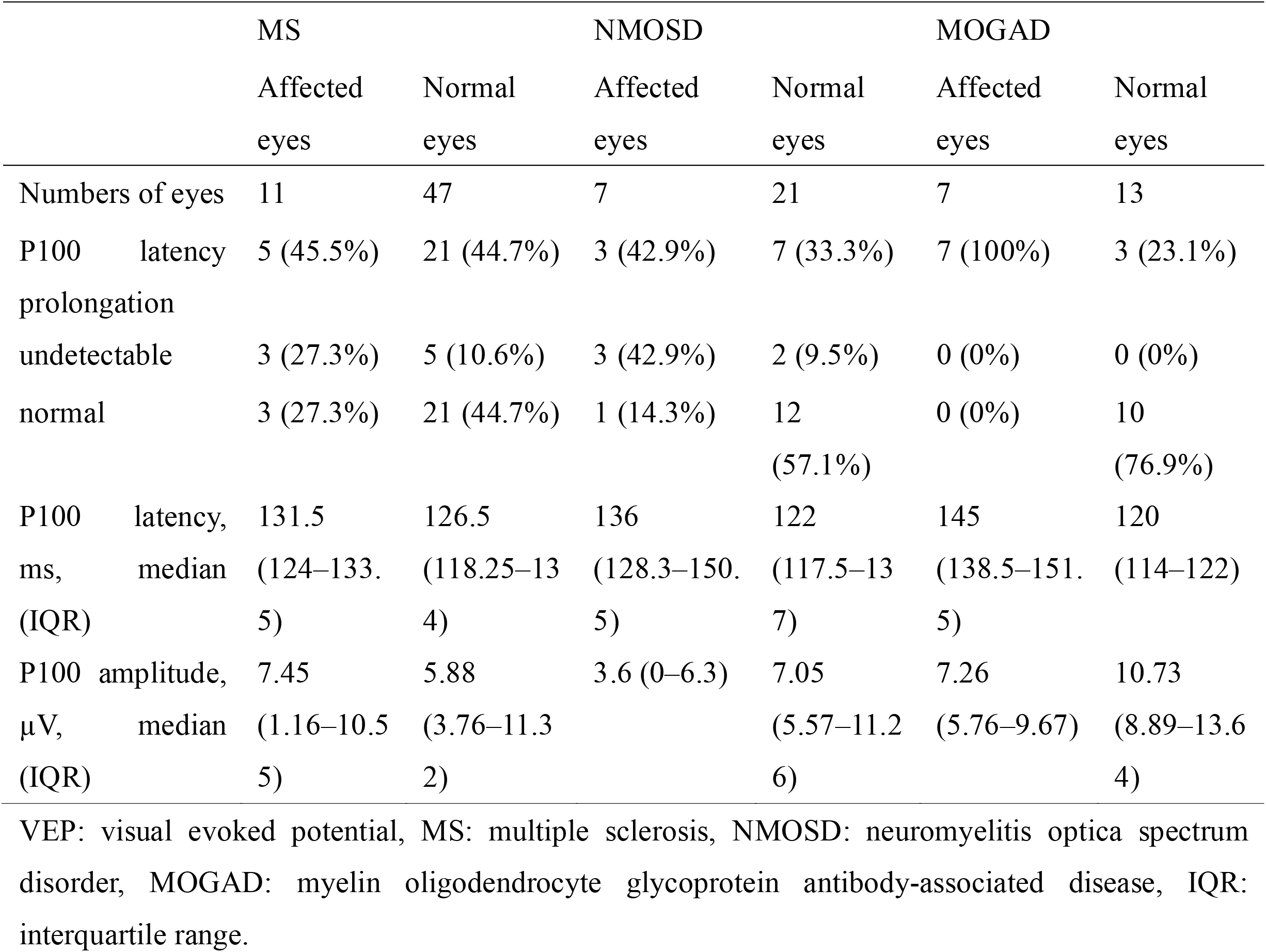
VEP in MS, NMOSD, and MOGAD.

**Table 3.**
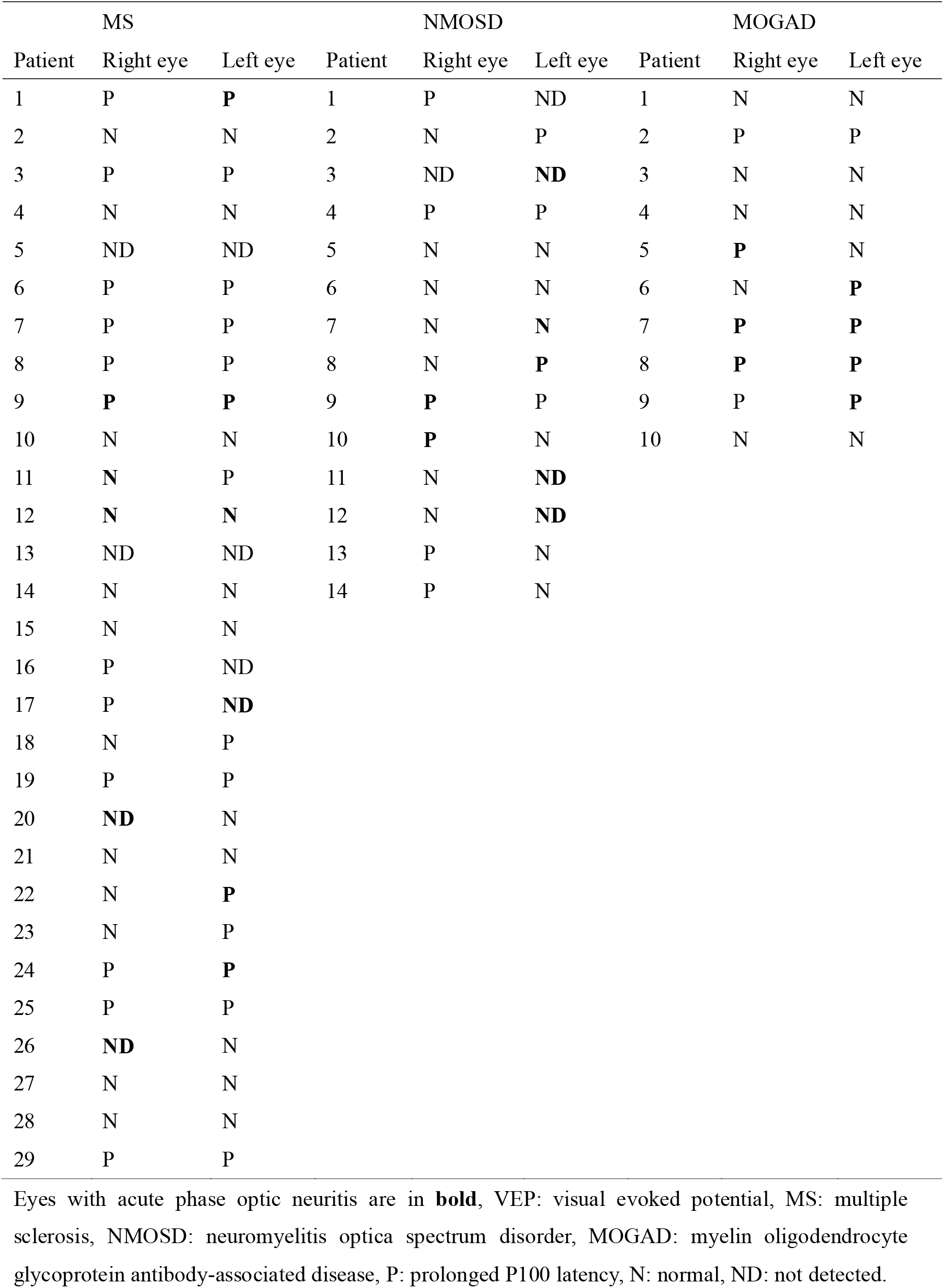
Individual VEP findings in patients with MS, NMOSD, and MOGAD.

**Figure 1.**
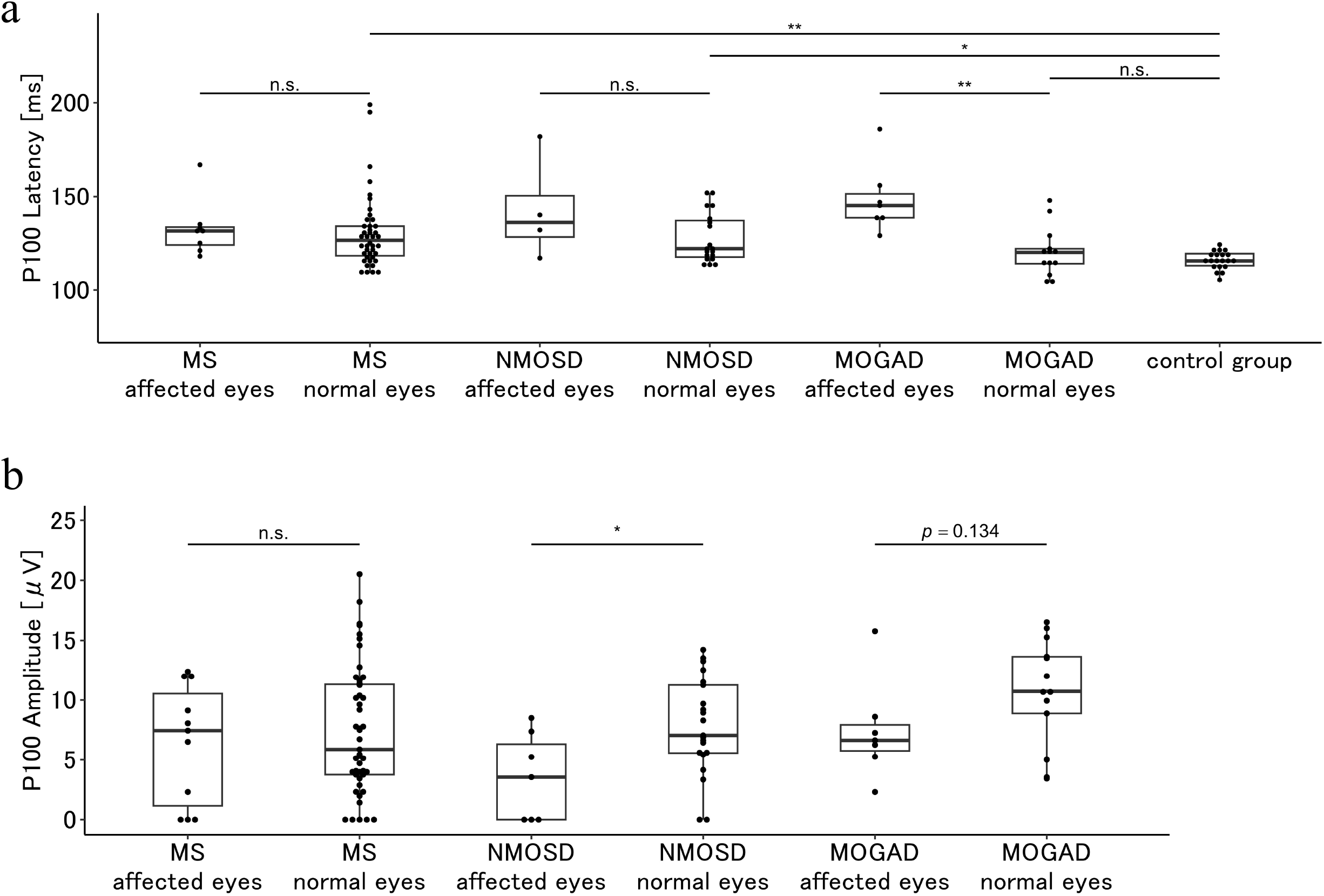
P100 latency and amplitude in MS, NMOSD, and MOGAD. (a) P100 latency is significantly longer in affected eyes than in normal eyes in patients with MOGAD; however, is not different between affected and normal eyes in patients with MS and NMOSD. P100 latency is significantly longer even in normal eyes (eyes without acute phase optic neuritis) in patients with MS and NMOSD compared to the control group. (b) P100 amplitude is significantly lower in affected eyes than in normal eyes in patients with NMOSD; however, is not different between affected and normal eyes in patients with MS and MOGAD. Significant differences are marked with * (p < 0.05) and ** (p < 0.001). MS: multiple sclerosis, NMOSD: neuromyelitis optica spectrum disorder, MOGAD: myelin oligodendrocyte glycoprotein antibody-associated disease, n.s.: not significant, affected eyes: eyes that were affected by acute phase optic neuritis, normal eyes: eyes without acute phase optic neuritis.

However, P100 latency was significantly prolonged in the eyes with acute phase ON than in the control group [131.5 ms (IQR, 124–133.5 ms) vs. 115.5 ms (IQR, 113.0–119.4 ms), *p <* 0.001]. Moreover, P100 latency was also significantly prolonged in the eyes without acute phase ON than in the control group [126.5 ms (IQR, 118.25–134.0 ms) vs. 115.5 ms (IQR, 113.0–119.4 ms), *p <* 0.001]. In addition, 7 of the 10 patients with unilateral ON, including those with acute phase ON or a prior history of ON, had abnormal VEP responses on the contralateral side.

### 3.2. Neuromyelitis optica spectrum disorder group

The NMOSD group comprised a total of 14 patients, who had a median age at disease onset of 50.5 years (IQR, 40.75–63.25 years). The median age at the time of VEP testing was 55 years (IQR, 46.25–64.25 years), and the disease duration ranged from 33 days to 42 years, with a median of 3.5 months (IQR, 2–19.75 months). At the time of VEP testing, patients’ EDSS median score was 4.5 (IQR, 2.75-5.0). Visual acuity assessments were available for 9 of 14 patients, with a median of 0.85. Of the 14 patients, 3 had cerebral lesions, 6 had brainstem lesions, and 12 had spinal cord lesions (Table S2). VEP testing was performed in 7 patients (6 women and 1 man) with acute phase ON and in 7 patients (4 women and 3 men) without acute phase ON, including those with a prior history of ON or without a history of ON (Table 1).

All NMOSD cases with acute phase ON had unilateral involvement, accounting for 7 eyes. Of the 7 eyes with acute phase ON, 3 had prolonged P100 latency, and 3 had undetectable VEP responses. In the remaining 21 eyes without acute phase ON, including 3 eyes with a prior history of ON and 18 eyes without a history of ON, 7 had prolonged P100 latency, and 2 had undetectable VEP responses (Table 2, 3). There were no significant differences in P100 latency between the eyes with acute phase ON and the other eyes (*p =* 0.33). P100 amplitudes were significantly lower in the eyes with acute phase ON, compared to those without acute phase ON [3.55 _μ_V (IQR, 0-6.31) vs. 7.05 (IQR, 5.57-11.26) *p =* 0.03]. P100 latency was significantly prolonged in the eyes with acute phase ON compared to the control group [136 ms (IQR, 128.25–150.5 ms) vs. 115.5 ms (IQR, 113.0–119.4 ms), *p =* 0.01]. Furthermore, P100 latency was significantly longer in the eyes without acute phase ON than in the control group [122 ms (IQR, 117.5–137 ms) vs. 115.5 ms (IQR, 113.0–119.4 ms), *p =* 0.003] (Figure 1).

### 3.3. Myelin oligodendrocyte glycoprotein antibody-associated disease group

The MOGAD group consisted of 10 patients who had a median age at disease onset of 32 years (IQR, 29.5–44.75 years). The median age at the time of VEP testing was 34.5 years (IQR, 29.5–45 years), and the disease duration ranged from 2 weeks to 5 years, with a median of 2 months (IQR, 1–52 months). At the time of VEP testing, patients’ EDSS median score was 3.5 (IQR, 2.0-4.0). Visual acuity assessments were available for 7 of 10 patients, with a median of 1.1. Of the 10 patients, 7 had cerebral lesions, 1 had a brainstem lesion, and 6 had spinal cord lesions (Table S3). VEP testing was performed in 5 patients (3 women and 2 men) with acute phase ON and in 5 patients (3 women, 2 men) without acute phase ON, including patients with a prior history of ON or without a history of ON (Table 1).

Among the MOGAD cases, acute phase ON was seen in 7 eyes (bilateral in 2 cases and unilateral in 3 cases). All 7 eyes with acute phase ON had detectable VEP responses and significantly longer P100 latency, compared with that in the other 13 eyes without acute phase ON, including 3 eyes with a prior history of ON and 10 eyes without a history of ON [145 ms (IQR, 138.5–151.5 ms) vs. 120 ms (IQR 114–122 ms), *p =* 0.004] (Table 2, 3) (Figure 1a). All 13 eyes without acute phase ON had detectable VEP responses, and of these, 10 exhibited normal VEP responses, including 3 eyes with a prior history of ON. P100 latency was prolonged in 3 eyes but was not significantly different between the eyes without acute phase ON and the control group [120 ms (IQR, 114–122 ms) vs. 115.5 ms (IQR, 113.0–119.4 ms), *p =* 0.43). P100 amplitude showed a tendency to be lower in eyes with acute phase ON than in eyes without ON [6.64 _μ_V (IQR, 5.76–7.935 _μ_V) vs. 10.73 _μ_V (IQR, 8.89–13.64 _μ_V), *p =* 0.134] (Figure 1b). Only one of the 6 cases with unilateral ON, including those with acute phase ON or a prior history of ON, had VEP abnormalities on the contralateral side.

## 4. Discussion

We presented detailed VEP data in patients with MS, NMOSD, and MOGAD. All patients with MOGAD had detectable VEP responses, regardless of the presence of acute phase ON. All eyes with acute phase ON had prolonged P100 latency, and the VEP response was abnormal in only 23% of eyes with nonacute phase ON. In contrast, VEP abnormalities were frequent in patients with MS and NMOSD, even in eyes without ON.

Previous studies on patients with chronic MOGAD reported that a history of ON did not affect P100 latency (Havla et al., 2021). Similarly, in our research, the VEP remained normal, even when there was a history of ON, provided there was no acute phase ON at the time of VEP testing. Conversely, even among eyes with nonacute phase ON secondary to MS, 55% had abnormal VEP results and significantly longer P100 latency, compared with that in the control group. Moreover, 70% of patients with MS who had unilateral ON exhibited abnormal VEP result on the contralateral side. Previous reports have also highlighted the existence of subclinical optic nerve dysfunction in a substantial percentage, ranging from 54% to 86%, of eyes without an ON attack (Grecescu, 2014; Sisto et al., 2005). These findings suggested that the subclinical damages to the optic nerve were fewer in MOGAD than in MS.

All patients with MOGAD exhibited detectable VEP responses and showed no statistically significant difference in P100 amplitudes, whether they had ON or not. However, among patients with acute phase ON in the NMOSD group, VEP responses were not detected in 42.9% of patients, which was consistent with previous reports on undetectable VEP responses in 47.4% of NMOSD cases (Neto et al., 2013). This difference in VEP patterns may reflect the different underlying pathological features. In NMOSD, the more pronounced secondary axonal degeneration secondary to astrocyte damage could explain the higher incidence of undetectable VEP responses (Wu et al., 2019). In MOGAD, the pathological change is more prone to demyelination than to axonal degeneration, because of the presence of MOG, which is the target antigen in MOGAD, in the myelin sheath of the central nervous system (Hoftberger et al., 2020). This may explain the relatively long P100 latency and highly detectable VEP responses in MOGAD cases. In the previous reports that suggested a reduction in P100 amplitude in MOGAD-associated ON, the patients included had relatively severe and protracted disease courses (Jarius et al., 2016). In contrast, most of our patients experienced ON for the first time or had relatively short disease durations.

Furthermore, among the patients with NMOSD in this study, P100 latency was significantly longer even in eyes without a history of ON than in the control group. This finding was consistent with that of a previous study, which indicated that patients with NMOSD may experience progressive prolongation of the P100 latency and a decrease in the P100 amplitude over time, even in the absence of clinical ON attacks (Ringelstein et al., 2020). This indicates that similar to MS, optic nerve dysfunction is possible in NMOSD, even without apparent ON.

The limitations of our study included the small sample size of the MOGAD group and the shorter disease duration of the MOGAD group than the MS and NMOSD groups. We plan to address this limitation by conducting long-term follow-up studies and gathering more comprehensive data in the future.

## 5. Conclusions

In MOGAD, VEP responses were detectable in all cases, P100 latency was prolonged in all eyes with acute phase ON, and abnormal VEP results were infrequent in the absence of clinical ON symptoms. Therefore, the presence of subclinical VEP abnormalities or undetectable VEP responses may differentiate MOGAD from MS and NMOSD. Given these characteristics, VEP testing may be valuable in the differential diagnosis of central nervous system inflammatory disorders.

## Data Availability

All data produced in the present work are contained in the manuscript and supplementary documents

## Author Contributions

Conceptualization, C.Y. and E.M.; methodology, T.N.; validation, M.A., S.N. and A.H.; data collection, A. Shigehisa., A. Sonoda.; writing – manuscript preparation, C.Y.; writing – review & editing, Y.H., K.M., A.H. and Y.S.; supervision, H.T. all authors have read and agreed to the published version of the manuscript.

## Institutional Review Board Statement

The study was conducted in accordance with the Declaration of Helsinki, and approved by the Institutional Review Board of Kagoshima University (protocol code 230141, approved on 12 October 2023).

## Informed Consent Statement

The requirement for written informed consent was waived by the institutional review board of Kagoshima University due to the retrospective nature of the study. Instead, information about the study was publicly disclosed on the institution’s website, allowing patients the opportunity to opt out of the research. This approach was approved by the institutional review board of Kagoshima University.

## Data Availability Statement

The original contributions presented in the study are included in the supplementary material, further inquiries can be directed to the corresponding author.

## Acknowledgments

We thank Dr. Takahashi Toshiyuki for the MOG antibody testing.

