## Supplementary for "Visual evoked potential in myelin oligodendrocyte glycoprotein antibody-associated disease"

Table S1. P100 Latency, amplitude, and presence/absence of optic neuritis in MS.

| Patient number | Eye | LP100 (ms) | AP100 (μV) | ON | prior ON | visual acuity | EDSS | Past attack |
| --- | --- | --- | --- | --- | --- | --- | --- | --- |
| 1 | Right | 166 | 5.44 |  |  | 0.7 | 3.5 | Brain |
|  | Left | 167 | 2.31 | + | + | 0.3 |  |  |
| 2 | Right | 113 | 3.75 |  |  | NA | 1 | Brainstem, Spine |
|  | Left | 116 | 4.06 |  | + | NA |  |  |
| 3 | Right | 128 | 5.88 |  |  | 1.2 | 2.5 | Brain, Brainstem, Spine |
|  | Left | 137 | 7.76 |  |  | 1.2 |  |  |
| 4 | Right | 123 | 16.27 |  |  | NA | 2 | Brain, Spine |
|  | Left | 125 | 20.52 |  |  | NA |  |  |
| 5 | Right | ND | ND |  |  | 0.7 | 7.5 | Spine |
|  | Left | ND | ND |  |  | 0.7 |  |  |
| 6 | Right | 128 | 7.51 |  |  | 1.2 | 4 | Brain, Brainstem |
|  | Left | 130 | 9.2 |  |  | 1.2 |  |  |
| 7 | Right | 199 | 4.08 |  | + | 0.7 | 1.5 | Brain, Spine |
|  | Left | 128 | 4.03 |  |  | 1 |  |  |
| 8 | Right | 138 | 4.01 |  |  | 1.2 | 6 | Brain, Brainstem, Spine |
|  | Left | 134 | 5.13 |  |  | 0.9 |  |  |
| 9 | Right | 133 | 6.51 | + |  | 1.2 | 2 | Brain, Spine |
|  | Left | 132 | 9.14 | + |  | 0.9 |  |  |
| 10 | Right | 117 | 11.64 |  |  | NA | 2.5 | Brain, Brainstem, Spine |
|  | Left | 109 | 12.76 |  |  | NA |  |  |
| 11 | Right | 125 | 12.02 | + |  | 1.2 | 3 | Brain |
|  | Left | 130 | 11.26 |  |  | 1.2 |  |  |
| 12 | Right | 121 | 11.95 | + |  | 1.2 | 2 | Brain, Spine |
|  | Left | 118 | 8.07 | + |  | 1.2 |  |  |
| 13 | Right | ND | ND |  |  | NA | 5 | Brain, Brainstem, Spine |
|  | Left | ND | ND |  |  | NA |  |  |
| 14 | Right | 113 | 10.2 |  |  | NA | 2 | Brain, Brainstem |
|  | Left | 116 | 11.89 |  |  | NA |  |  |
| 15 | Right | 109 | 3.44 |  |  | NA | 4 | Brain, Spine |
|  | Left | 110 | 3.76 |  |  | NA |  |  |
| 16 | Right | 158 | 3.94 |  |  | NA | 2 | Brain, Brainstem, Spine |
|  | Left | ND | ND |  |  | NA |  |  |
| 17 | Right | 129 | 9.64 |  |  | 1.2 | 5 | Brain |
|  | Left | ND | ND | + |  | 0.01 |  |  |
| 18 | Right | 124 | 7.82 |  |  | 0.7 | 4 | Brain, Brainstem, Spine |
|  | Left | 143 | 7.64 |  |  | 1 |  |  |
| 19 | Right | 151 | 2.31 |  | + | NA | 1.5 | Brain, Brainstem, Spine |
|  | Left | 132 | 2.88 |  | + | NA |  |  |
| 20 | Right | ND | ND | + |  | 0.01 | 5 | Brain |
|  | Left | 123 | 5.32 |  |  | 0.6 |  |  |
| 12 | Right | 119 | 15.15 |  |  | NA | 6 | Brain, Brainstem, Spine |
|  | Left | 115 | 16.39 |  |  | NA |  |  |
| 22 | Right | 121 | 14.58 |  | + | 1.2 | 1 | Brain |
|  | Left | 131 | 7.45 | + | + | 1.2 |  |  |
| 23 | Right | 123 | 1.42 |  |  | 1 | 4 | Brain |
|  | Left | 134 | 1.98 |  |  | 1 |  |  |
| 24 | Right | 131 | 15.52 |  |  | 1.2 | 5 | Brain |
|  | Left | 135 | 12.39 | + |  | fingers |  |  |
| 25 | Right | 140 | 4.71 |  |  | 1.2 | 2 | Brain, Spine |
|  | Left | 195 | 2.33 |  | + | 0.5 |  |  |
| 26 | Right | ND | ND | + |  | 0.01 | 2 | Brain, Brainstem, Spine |
|  | Left | 110 | 11.38 |  |  | 1.2 |  |  |
| 27 | Right | 119 | 6.73 |  |  | NA | 2.5 | Brain, Spine |
|  | Left | 120 | 5.13 |  |  | NA |  |  |
| 28 | Right | 122 | 10.16 |  |  | NA | 4.5 | Brain, Brainstem, Spine |
|  | Left | 118 | 10.39 |  |  | NA |  |  |
| 29 | Right | 134 | 18.21 |  |  | NA | 7.5 | Brain, Spine |
|  | Left | 149 | 11.95 |  | + | NA |  |  |

MS: multiple sclerosis, ON: acute phase optic neuritis, prior ON: prior optic neuritis, EDSS: Expanded Disability Status Scale, Past attack: lesions other than optic nerve confirmed clinically or radiologically, LP100 (ms): P100 wave latency in milliseconds, AP100 (µV): P100 wave amplitude in microvolts, ND: not detected, NA: not available.

Table S2. P100 Latency, amplitude, and presence/absence of optic neuritis in NMOSD.

| Patient number | Eye | LP100 (ms) | AP100 (μV) | ON | prior ON | visual acuity | EDSS | Past attack |
| --- | --- | --- | --- | --- | --- | --- | --- | --- |
| 1 | Right | 145 | 11.51 |  |  | 0.5 | 7 | Spine |
|  | Left | ND | ND |  | + | 0.03 |  |  |
| 2 | Right | 113 | 13.26 |  |  | 1 | 0 |  |
|  | Left | 152 | 13.52 |  | + | 1.2 |  |  |
| 3 | Right | ND | ND |  |  | 1.2 | 4 | Brain, Spine |
|  | Left | ND | ND | + | + | 1 |  |  |
| 4 | Right | 145 | 3.35 |  |  | 0.7 | 6.5 | Brainstem, Spine |
|  | Left | 152 | 4.15 |  |  | 1.2 |  |  |
| 5 | Right | 119 | 11.26 |  |  | NA | 3.5 | Brainstem, Spine |
|  | Left | 118 | 12.52 |  |  | NA |  |  |
| 6 | Right | 121 | 6.89 |  |  | NA | 2 | Spine |
|  | Left | 120 | 5.64 |  |  | NA |  |  |
| 7 | Right | 117 | 5.57 |  |  | NA | 1 | Brainstem, Spine |
|  | Left | 117 | 8.51 | + |  | NA |  |  |
| 8 | Right | 124 | 14.22 |  |  | 1.2 | 5 | Brainstem, Spine |
|  | Left | 182 | 3.55 | + |  | 0.03 |  |  |
| 9 | Right | 140 | 7.38 | + |  | 0.6 | 7.5 | Brainstem, Spine |
|  | Left | 138 | 6.63 |  |  | 0.6 |  |  |
| 10 | Right | 132 | 5.24 | + |  | NA | 2.5 | Spine |
|  | Left | 114 | 6.44 |  |  | NA |  |  |
| 11 | Right | 114 | 7.05 |  |  | 1.2 | 5 | Brain, Brainstem, Spine |
|  | Left | ND | ND | + |  | 0.04 |  |  |
| 12 | Right | 116 | 9.7 |  |  | 1.2 | 5 |  |
|  | Left | ND | ND | + |  | 0.03 |  |  |
| 13 | Right | 134 | 8.3 |  |  | NA | 2 | Brain, Spine |
|  | Left | 122 | 8.95 |  |  | NA |  |  |
| 14 | Right | 136 | 5.43 |  | + | 0.9 | 4 | Spine |
|  | Left | 122 | 9.2 |  |  | 1.2 |  |  |

NMOSD: neuromyelitis optica spectrum disorder, ON: acute phase optic neuritis, prior ON: prior optic neuritis, EDSS: Expanded Disability Status Scale, Past attack: lesions other than optic nerve confirmed clinically or radiologically, LP100 (ms): P100 wave latency in milliseconds, AP100 (µV): P100 wave amplitude in microvolts, ND: not detected, NA: not available.

Table S3. P100 latency, amplitude, and presence/absence of optic neuritis in MOGAD.

| Patient number | Eye | LP100 (ms) | AP100 (μV) | ON | prior ON | visual acuity | EDSS | Past attack |
| --- | --- | --- | --- | --- | --- | --- | --- | --- |
| 1 | Right | 105 | 13.64 |  |  | NA | 4 | Brain, Spine |
|  | Left | 104 | 16.52 |  |  | NA |  |  |
| 4 | Right | 142 | 8.89 |  |  | 1.2 | 4.5 | Brain |
|  | Left | 148 | 5.01 |  |  | 1.2 |  |  |
| 2 | Right | 114 | 15.26 |  |  | NA | 7 | Brain, Spine |
|  | Left | 115 | 13.52 |  | + | NA |  |  |
| 3 | Right | 122 | 12.01 |  |  | 1.5 | 4 | Brain, Spine |
|  | Left | 121 | 10.64 |  | + | 0.7 |  |  |
| 5 | Right | 129 | 6.26 | + | 2 | 1 | 1 |  |
|  | Left | 115 | 9.95 |  |  | 1.5 |  |  |
| 6 | Right | 108 | 16.02 |  |  | 1.5 | 4 | Brain, Spine |
|  | Left | 147 | 5.26 | + |  | 0.6 |  |  |
| 7 | Right | 138 | 2.31 | + |  | 0.5 | 3 | Brain, Spine |
|  | Left | 139 | 15.77 | + |  | 1.5 |  |  |
| 8 | Right | 145 | 8.61 | + |  | 1 | 1.5 | Brainstem |
|  | Left | 156 | 7.26 | + |  | 0.9 |  |  |
| 9 | Right | 129 | 10.73 |  |  | 1.2 | 2 |  |
|  | Left | 186 | 6.64 | + |  | 0.01 |  |  |
| 10 | Right | 120 | 3.54 |  | + | NA | 2 | Brain, Spine |
|  | Left | 121 | 3.43 |  |  | NA |  |  |

MOGAD: myelin oligodendrocyte glycoprotein antibody-associated disease, ON: acute phase optic neuritis, prior ON: prior optic neuritis, EDSS: Expanded Disability Status Scale, Past attack: lesions other than optic nerve confirmed clinically or radiologically, LP100 (ms): P100 wave latency in milliseconds, AP100 (µV): P100 wave amplitude in microvolts, NA: not available.
